# Diagnostic accuracy of point-of-care testing (POCT) devices for postpartum haemorrhage: A protocol of a systematic review and meta-analysis

**DOI:** 10.1101/2023.06.11.23291255

**Authors:** Eishin Nakamura, Takahiro Mihara, Yuriko Kondo, Hisashi Noma, Sayuri Shimizu

## Abstract

**Introduction:** In postpartum haemorrhage (PPH), coagulation factors, mainly fibrinogen, are exhausted early during the course of the disease, which can easily lead to disseminated intravascular coagulation (DIC) and cause maternal mortality. Although some studies have reported the efficacy of point-of-care testing (POCT) devices in detecting hypofibrinogenaemia, little research has been conducted on their usefulness in the diagnosis of PPH. We plan to perform a systematic review of the diagnostic accuracy of existing POCT devices for the diagnosis of hypofibrinogenaemia in PPH.

**Methods and analysis:** We plan to identify studies using POCT devices for PPH and perform a systematic review and meta-analysis of their accuracy of diagnosing hypofibrinogenaemia. The results of fibrinogen measurements using the conventional Clauss method are used as reference standards. Literature searches will be conducted using electronic databases (MEDLINE, Embase, Cochrane Database, and Web of Science), and two reviewers will screen independently from the search results. The eligible literature will be used to synthesise true positive, false positive, true negative, and false negative results for the diagnosis of hypofibrinogenaemia. We will estimate the diagnostic accuracy of POCT devices for diagnosing hypofibrinogenaemia in patients with PPH using the Reitsma-type bivariate random-effects synthesis model and the hierarchical summary receiver operating characteristic curve.

**Ethics and dissemination:** This systematic review will be conducted through the secondary use of literature extracted from electronic databases. There are no ethical issues associated with this research. The final integrated results will be submitted to a peer-reviewed journal.

**Protocol registration:** The study protocol was registered with University Hospital Medical Information Network Clinical Trials (UMIN000048272) and PROSPERO (CRD42023394785).

**Strengths and limitations of the study:**

- Studies on the usefulness of POCT devices in the diagnosis of PPH are limited. Our study will perform a systematic review of the diagnostic accuracy of existing POCT devices for the diagnosis of hypofibrinogenaemia in PPH.
- The study will estimate the diagnostic accuracy of POCT devices using the Reitsma-type bivariate random-effects synthesis model and the hierarchical summary receiver operating characteristic curve.
- The threshold for diagnosing hypofibrinogenaemia, the definition of PPH, and the POCT devices used may vary between studies and could be a potential source of heterogeneity. Since most primary studies are observational, it is expected that many unpublished studies will exist.
- The applicability of the study results may be limited since this systematic review only pertains to the use of POCT devices in pregnant patients with PPH. The number of studies may be limited since there is a wide variety of POCT devices used in PPH.

## INTRODUCTION

### Background

Postpartum haemorrhage (PPH) causes massive blood loss during labour and is one of the leading causes of maternal death.(1) PPH often presents with severe coagulopathy complicated by disseminated intravascular coagulation (DIC). The level of fibrinogen in the blood, which is considered the first to fall below the haemostatic threshold is particularly important, and blood fibrinogen levels correlate with the severity of PPH.(2) Early diagnosis of hypofibrinogenaemia and the initiation of transfusion therapy are important for improving maternal outcomes.

Conventional fibrinogen testing using the Clauss method is time-consuming and expensive to be installed in primary medical facilities. In recent years, point-of-care testing (POCT) devices that simply estimate blood coagulation function have been used as an initial treatment for PPH, and there are reports of their usefulness in PPH.(3) Moreover, the POCT device allows for rapid testing at a low cost, and the test results correlate well with fibrinogen levels in conventional blood tests.(4) Although many reports on the effectiveness of POCT devices in clinical practice have been recognised in the fields of cardiovascular surgery(5) and emergency medicine(6), there are limited reports on their use in PPH. Since PPH has a different mechanism than other haemorrhagic diseases such as trauma or cardiac surgery, the usefulness of POCT in other diseases may not be directly applicable.

Early diagnosis of coagulopathy, especially hypofibrinogenaemia, in patients with PPH, and appropriate transfusion therapy may contribute to decreased total blood loss and mortality. Hence, early diagnosis of coagulopathy using POCT devices is important for improving the prognosis of PPH.

### Clinical role of the index test

In clinical practice, dry haematology, thromboelastography, and thromboelastometry are commonly used POCT devices.

Dry haematology can directly and rapidly measure fibrinogen by measuring the viscosity of the magnetic particle motion signal using scattered light and calculating the fibrinogen value from the absorbance.(7) Therefore, it is reasonable to use it for the diagnosis of PPH, in which fibrinogen levels are particularly important, among other coagulation factors.

Thromboelastography enables the comprehensive evaluation of coagulation and haemostatic function, including the effect of platelets, using whole blood samples. For TEG6s®, the parameters that could be measured as actual values are 1. reaction time (R), 2. Kinetics (K), 3. Angle, 4. maximum amplitude (MA), 5. whole blood clot lysis index at 30 minutes after MA(Ly30), 6. amplitude 10(A-10) and 7. Functional fibrinogen (FLEV), 8. Activated clotting time (ACT) (8). TEG6s can simultaneously measure four different reagents: Citrated Kaolin (CK), Citrated Kaolin with Heparinase (CKH), Citrated Rapid TEG (CRT), Citrated Functional Fibrinogen (CFF), and Citrated Rapid TEG (CRT).

The measurement principle of thromboelastometry is similar to that of thromboelastography but requires a large number of reagents and pipetting steps. (9) Thromboelastography and thromboelastometry often investigate and report parameters that correlate more with fibrinogen values since direct fibrinogen measurement is not possible.(10)

### Objectives

The Population, Index tests, and Target condition (PIT) framework(11) in this study is shown in Table 1.(12)

**Table 1:** Population, Index tests, Target condition (PIT) framework

|  |  |
| --- | --- |
| Population | Pregnant patients after 20 weeks gestation and postpartum patients. |
| Index tests | The following POCT devices are defined as index tests.<br>1. Dry haematology<br>2. Thromboelastography<br>3. Thromboelastometry |
| Target condition | Hypofibrinogenemia in patients with PPH |

POCT, point-of-care testing; PPH, postpartum haemorrhage.

The purpose of this study is to perform a systematic review of the diagnostic accuracy of POCT devices in diagnosing hypofibrinogenaemia in patients with PPH. The population, index test, and target conditions are presented in Table 1. The reference standard for diagnosing hypofibrinogenaemia will be based on fibrinogen levels measured using the conventional Clauss method, and the cut-off value for diagnosing hypofibrinogenaemia will be determined based on the values set in each individual study. Figure 1 shows the clinical flowchart of the included studies.

**Figure 1:**
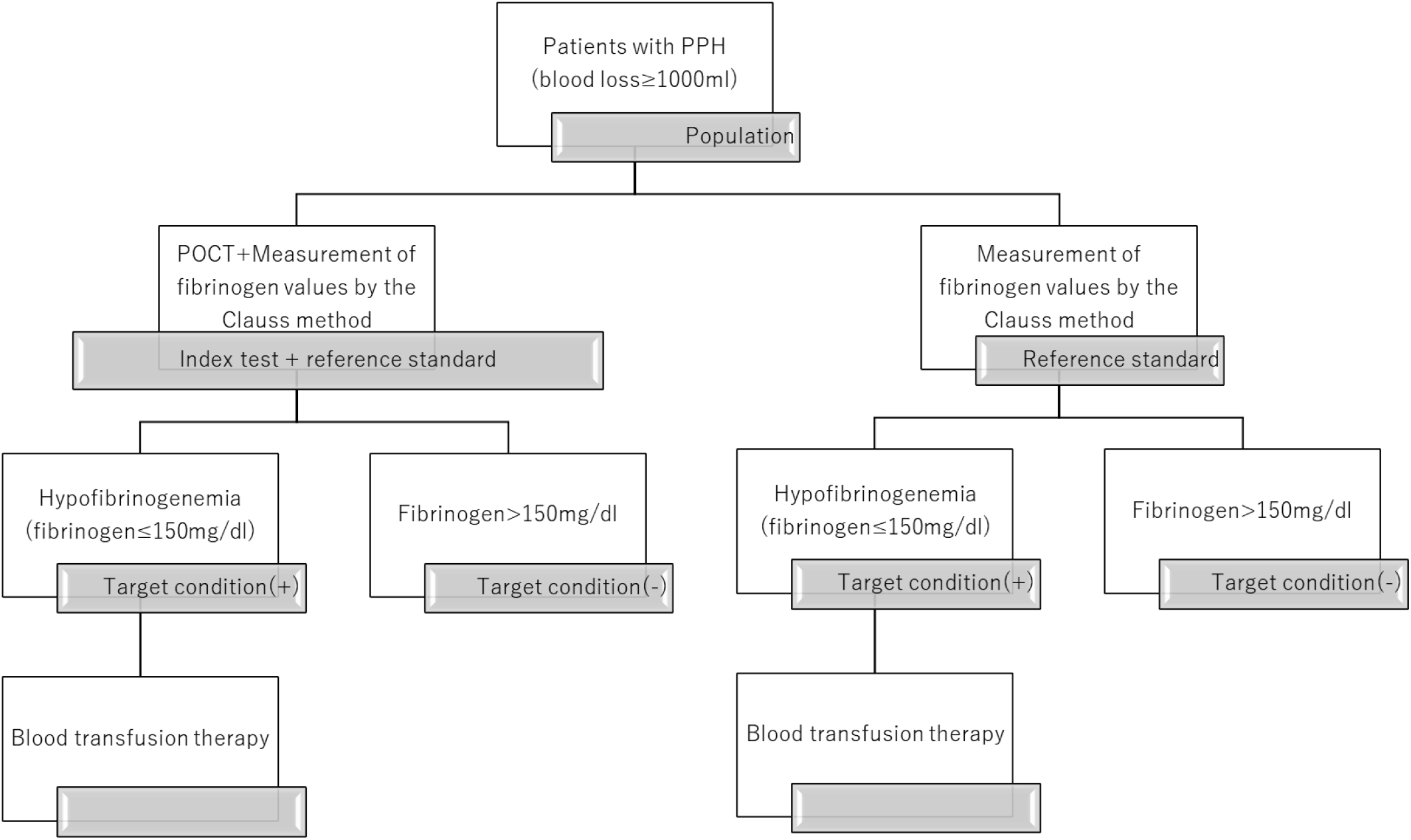
clinical flowchart of postpartum haemorrhage (PPH) patients

### Significance of this review

As mentioned above, there have been numerous reports on the usefulness of POCT devices in emergency medicine and cardiovascular surgery. However, there are few reports on their use in PPH. Compared to non-pregnant women, pregnant women are characterised by circulatory dynamics and blood coagulability. In pregnant women, the circulating blood volume is increased by up to 50%, all coagulation factor activities except for factor 13 are increased, and the fibrinolytic system is relatively suppressed.(12,13) Since fibrinogen requires the highest concentration of any coagulation factor during haemostasis, high-volume transfusion in the setting of PPH is easily complicated by dilutional coagulopathy.(14,15) In addition, PPH consumes large amounts of coagulation factors to stop bleeding from the placental abruption site, which may result in consumptive coagulopathy.(16) Since fibrinogen is the first to be consumed in these coagulation disorders, DIC progresses more rapidly in PPH than in other haemorrhagic diseases. Therefore, given the unique aspects of pregnancy, it is necessary to investigate the effectiveness of POCT devices separately in patients with PPH.

If POCT devices demonstrate high diagnostic accuracy for detecting hypofibrinogenaemia in PPH, they can be used to guide treatment strategies for the early identification of coagulation disorders and appropriate transfusion therapy.

## METHODS AND ANALYSIS

This systematic review and meta-analysis follows the Preferred Reporting Items for Systematic Reviews and Meta-Analyses (PRISMA) statement and the PRISMA-Diagnostic Test Accuracy (DTA) statement.(17,18) The study protocol was registered with University Hospital Medical Information Network Clinical Trials (UMIN000048272) and PROSPERO (CRD42023394785).

### Eligibility criteria

#### Participants

Pregnant women and postpartum women after 20 weeks gestation and patients with blood loss >1000 ml will be included. To examine the effectiveness of POCT devices in investigating the parameters of bleeding in pregnant women, studies that only include pregnant women with congenital coagulopathy will be excluded.

#### Setting

Studies in which the initial treatment for PPH or follow-up after transfusion was administered with POCT will be included. Pilot studies conducted on healthy pregnant women and postpartum mothers will be excluded.

#### index test

Only studies using Dry haematology (e.g. CG02N®), Thromboelastography (e.g. TEG®), or Thromboelastometry (e.g. ROTEM®) as an index test will be included.

#### reference standard

Only studies that use the Clauss method for fibrinogen measurement as the reference standard will be included. There are three methods for measuring fibrinogen in blood: the Clauss method, the salt precipitation method, and the immunoassay method. The Clauss method is the most widely used internationally, and studies using salting-out and immunoassay methods are excluded.(19) A low blood fibrinogen level in pregnant women is reported to be a risk for PPH at ≤200 mg/dl and a risk for requiring massive blood transfusions at ≤130–155 mg/dl.(20–22) The definition of hypofibrinogenaemia is inconsistent in the literature with fibrinogen ≤ 150 mg/dl and ≤ 200 mg/dl.(23) We plan to collect the literature regardless of the cut-off value of hypofibrinogenaemia and integrate the results, and we will conduct a subgroup analysis according to the cut-off value if there are a sufficient number of studies (i.e., ≥ five studies) for each cut-off value.

#### target condition

This study will include only PPH patients diagnosed with hypofibrinogenemia.

#### Types of studies

Only studies involving humans will be included, and literature on animals will be excluded. Studies will include randomised controlled trials (RCTs) and non-RCTs (non-randomised controlled trials, split time-series analyses, before/after comparisons, cohort studies, and case-control studies). Non-published studies (e.g. conference abstracts and clinical trial protocols) will also be included.

### Information sources and search strategy

The literature will be searched using electronic databases such as MEDLINE, Embase, the Cochrane Database, and Web of Science, and references to the corresponding literature will also be evaluated. A literature search of these databases will be conducted on 15/06/2023.

The search formula for PubMed is shown below in Table 2.

**Table 2:** The search strategy for PubMed

| Search Number | Query |
| --- | --- |
| 1 | "postpartum hemorrhage"[MeSH Terms] OR (postpartum[tiab] AND hemorrhage[tiab]) OR (postpartum[tiab] AND haemorrhage[tiab]) |
| 2 | "point of care testing"[MeSH Terms] OR "Point-Of-Care"[tiab] OR (point*[tiab] AND care[tiab] AND test*[tiab]) |
| 3 | dry[tiab] AND hematology[tiab] |
| 4 | "thrombelastography"[MeSH Terms] OR "Thromboelastometry"[tiab] |
| 5 | "blood coagulation tests"[MeSH Terms] OR (blood[tiab] AND coagulation[tiab] AND test*[tiab]) |
| 6 | "viscoelasti*" [tiab] AND ("haemostat*" [tiab] OR "hemostatics"[MeSH Terms] OR "hemostasis"[MeSH Terms] ) |
| 7 | #2 OR #3 OR #4 OR #5 OR #6 OR #7 |
| 8 | #1 AND #7 |

### Study selection

#### Data management and selection process

The list of references obtained from the electronic databases will be scanned using literature management software (EndNote®) to remove duplicate references. The titles and abstracts will be screened by two independent reviewers (EN and YK) to assess the inclusion criteria. For references determined to be eligible by at least one of the two reviewers in the primary screening, we will proceed to the secondary screening process using the full text. Secondary screening using the full text will be conducted for references that qualify in the primary screening and for those that cannot be determined to be eligible based on the title and abstract. Disagreements regarding the eligibility for the study will be resolved through discussion.

#### Data collection process

Two independent reviewers (EN and YK) will extract data from the eligible literature. If there are discrepancies in data extraction between the two parties regarding the extraction of data, they will be resolved through discussion.

#### Data items

Maternal age at delivery, weeks of delivery, primiparity, maternal body mass index, method of delivery (vaginal or cesarean), cause of PPH (atonic bleeding, perineal laceration, retained or adhered placenta, placental abruption, amniotic fluid embolism), total blood loss, total blood transfusion requirements (Red Cell Concentrate, Fresh Frozen Plasma, Platelet Concentrate, fibrinogen concentrate), maternal deaths, blood fibrinogen levels measured by the Clauss method at the time of POCT device use, type of POCT device used, measurement results of POCT devices [(1) dry-haematology: fibrinogen, (2) thromboelastography: R, K, MA, Ly30, A-10, FLEV, (3) thromboelastometry: CT, CF, CFT, alpha, A10, MCF, LI30, LI60, ML], and the number of patients in the true positive, false positive, true negative, and false positive will be collected as diagnostic accuracy for hypofibrinogenemia.

### Outcomes and prioritization

#### Primary outcome

The primary outcome is the diagnostic accuracy of the POCT device to determine hypofibrinogenemia. Hypofibrinogenemia is diagnosed based on the fibrinogen values measured using the Clauss method.

#### Secondary outcomes

The secondary outcome is the correlation coefficient between the fibrinogen value measured using the Clauss method and the test values measured using POCT.

### Risk of bias in individual studies

QUADAS-2 (24) will be used to assess the risk of bias for each study; QUADAS-2 is classified into the following four domains: (1) participant selection, (2) index test, (3) reference standard, and (4) flow and timing. For each domain, the risk of bias is classified as ‘low’, ‘unclear’, and ‘high’, and concerns about applicability are also classified as ‘low’, ‘unclear’, and ‘high’. Signalling questions are also included to facilitate the evaluation of the risk of bias for each domain.

### Data synthesis and meta-analysis

Firstly, we will present the predictive accuracy measures of individual studies based on the reported counts of true positives, false positives, false negatives, and true negatives; sensitivities and specificities will be calculated, accompanied by their corresponding 95% confidence intervals. Coupled forest plots will be used to visualize sensitivities and specificities of POCT for diagnosing hypofibrinogenaemia. In addition, scatter plots will be generated for pairs of sensitivities and specificities to perform exploratory evaluations and assess heterogeneity.

Secondly, we will perform synthesis analyses using Reitsma’s bivariate random-effects model to account for possible heterogeneities across studies based on study-specific sensitivities and specificities.(25) The results will be the summary estimates of sensitivities and specificities, along with their heterogeneity variances (τ^2^). Furthermore, we will generate summary receiver operating characteristic (SROC) curves(26) based on bivariate random-effects model estimates and report the areas under the curves (AUCs) as a summary measure of diagnostic accuracy.(27,28) For statistical inference, we will employ standard restricted maximum likelihood estimation for Reitsma’s model(25) and use the bootstrap method to calculate 95% confidence intervals for the AUCs of the SROC curves.(29) We will also perform a generalised Egger test for multivariate meta-analysis to assess potential publication bias.(30) Statistical analyses will be performed using R software (R Development Core Team, Vienna, Austria) and RStudio (RStudio, Boston, Massachusetts, USA).

### Confidence of cumulative evidence

Once sufficient results have been obtained, the Grading of Recommendations, Assessment, Development, and Evaluations (GRADE) will be used to evaluate the certainty of evidence.(31)

### Subgroup analyses

If different thresholds for hypofibrinogenaemia are adopted among the eligible studies, subgroup analyses will be considered according to the prespecified clinically relevant thresholds (≤150 mg/dl, ≤200 mg/dl). In addition, to evaluate possible heterogeneity, subgroup analyses will be performed based on the types of POCT devices used (dry haematology, thromboelastography, and thromboelastometry).

## DISCUSSION

Early detection of hypofibrinogenaemia in PPH and appropriate coagulation factor replacement are crucial to improve the prognosis of PPH. However, no previous systematic review has reported the usefulness of POCT devices in PPH. This systematic review focuses on the diagnostic accuracy of POCT for hypofibrinogenaemia and may provide data for the effective use of POCT devices in PPH. This study will focus on blood fibrinogen levels in patients with obstetric haemorrhage. Fibrinogen level is the most important coagulation index in the blood of patients with PPH. Although many coagulation factors are necessary for haemostasis, fibrinogen is required in the highest concentration in the blood.(32) When massive haemorrhage occurs due to PPH, high-volume intravenous infusion to maintain blood pressure can easily lead to dilutional coagulopathy.(33) Furthermore, there is a risk of consumptive coagulopathy, in which a large amount of coagulation factors is consumed to stop bleeding from the placental abruption surface. Moreover, fibrinogen is also the first to be consumed.(34) Therefore, it is reasonable to assume that early recognition of hypofibrinogenemia and appropriate blood transfusion therapy will improve the prognosis of PPH compared to other massive haemorrhages.

In PPH, early identification of hypofibrinogenaemia and administering fibrinogen concentrate may reduce the total amount of blood transfusion required and improve the prognosis.(23) Early diagnosis of hypofibrinogenaemia using POCT devices and the development of transfusion protocols may improve maternal outcomes in PPH and conserve medical resources, including transfusion products.(35) This systematic review aims to determine the effectiveness of POCT in the diagnosis of hypofibrinogenaemia in patients with PPH. If POCT devices are reliable for accurately diagnosing hypofibrinogenaemia and determining the minimum transfusion volume needed to correct coagulopathy, they could potentially increase maternal survival rates and reduce the burden on medical resources.

### Limitations and implications

This systematic review has several limitations. First, the heterogeneity among the studies may be an issue. The threshold for diagnosing hypofibrinogenaemia, the definition of PPH, and the POCT devices used may vary between studies and could be a potential source of heterogeneity. Second, since most primary studies are observational, it is expected that many unpublished studies will exist. Third, the applicability of the study results may be limited since this systematic review only pertains to the use of POCT devices in pregnant patients with PPH. Fourth, the number of studies may be limited since there is a wide variety of POCT devices used in PPH. If there are insufficient reports for each device, the results of all POCT devices may be combined, making it difficult to determine the most effective device since there will be no direct comparison between the devices.

## Data Availability

All data produced in the present work are contained in the manuscript

## ETHICS AND DISSEMINATION

This systematic review will finally be submitted for publication in a peer-reviewed journal. The data used in this study do not include individual patient data, so there are no patient privacy concerns. There are also no restrictions on the release of data. Any significant changes from this protocol will be described in detail.

## Acknowledgements

We would like to thank Editage (www.editage.com) for English-language editing.

## Author contributions

Conceptualization: Ehisin Nakamura, Takahiro Mihara, Sayuri Shimizu

Methodology: Eishin Nakamura, Takahiro Mihara, Yuriko Kondo, Hisashi Noma

Supervision: Takahiro Mihara, Sayuri Shimizu, Hisashi Noma

Writing – Original Draft Preparation: Eishin Nakamura

Writing – Review & Editing: Eishin Nakamura, Takahiro Mihara, Yuriko Kondo, Hisashi Noma, Sayuri Shimizu.

